# NETosis is an important component of chronic inflammation in patients with heart failure

**DOI:** 10.1101/2024.07.09.24310187

**Authors:** Sawa Kostin, Manfred Richter, Florian Krizanic, Benjamin Sasko, Theodoros Kelesidis, Nikolaos Pagonas

## Abstract

**Background and aim:** We have previously demonstrated that heart failure (HF) is characterized by low-grade myocardial inflammation. However, the role of neutrophils (N), neutrophil extracellular traps (NETs) and neutrophil cell death by NETosis in the myocardium of patients with HF remains largely unknown. The present study investigated the number of neutrophils (N) and their proportion undergoing NETosis and developing NETs in HF.

**Methods:** We used quantitative confocal microscopy and NETosis markers in the left ventricular biopsies obtained from 5 control and from patients with HF due to dilated (DCM, n=7), inflammatory (infCMP, n=7) and ischemic cardiomyopathy (ICM, n=7). We used immunolabeling for CD45, CD66b and CD11b for (N) and citrullinated histone3 (citH3), peptidylarginine deiminase-4 (PAD-4), neutrophil elastase (NE) and myeoloperoxidase (MPO) for NETosis. These proteins were also investigated by quantitative fluorescence intensity analysis, Western blot and quantitative polymerase chain reaction (qPCR).

**Results:** Compared to control, the number of N was increased 3-4 times in HF. We found that using a single marker for NETosemarkers, 43.2% of N in DCM, 46.7% in ICM and 57.3% in infCMP experienced NETosis. The use of double labeling (NE with CitH3) showed that 55.6% of N developed NETosis in DCM, 57.9% in ICM and 79.4% in infCMP. The difference between the N who underwent NETosis in infCMP and those in DCM was statistically different (p<0.01). The proportion of N who developed NETosis or formed NETs in control tissue was less than 5% and differed significantly from that in HF patients, regardless of etiology (p<0.01). These results were confirmed by quantitative fluorescence analysis, Western blot and qPCR.

**Conclusions:** This is the first study to show the occurrence of NETosis in human hearts *in situ* indicating that NETosis is an important component of low-grade myocardial inflammation in HF.

**What is new?:** Low-grade myocardial inflammation is a typical feature of heart failure and neutrophil cell death (NETosis) is an important component of this pathological process.

**What are the clinical implications?:** Preventing excessive neutrophil activation and inhibiting the major components involved in the NETosis program (neutrophil elastase, myeloperoxidase and peptidylarginine deiminase-4) are perspective targets for the treatment of HF.

## Introduction

The hallmark of heart failure (HF) is pathological structural myocardial remodeling including cardiomyocyte hypertrophy, fibrosis, cardiomyocyte cell death and fibrosis. Chronic, low-grade myocardial inflammation is also one of the major factors implicated the development and progression of HF [1–3], however most of the clinical trials of anti-cytokine therapy have shown modest results, indicating that we still have to further study the mechanisms of inflammation in HF [1, 2].

Polymorphonuclear neutrophils are terminally differentiated cells and are the most abundant subset of leukocytes and the major constituents of the innate immune system. These short-lived cells are powerful inducers of the inflammatory response, One of the key function of neutrophils is the formation of the extracellular web-like network defined as NET (Neutrophil Extracellular Trap). Originally, this unique process of NETs formation was linked to neutrophil cell death and was therefore called NETosis [8]. Originally, this unique process of NETs formation was linked to neutrophil cell death and was therefore called NETosis [3]. However, NETs formation can either occur with preservation of neutrophilic functions (vital NETosis) or with neutrophil cell death (suicidal or lytic NETosis).

NETs formation and NETosis require neutrophil activation of the arginine deiminase 4 (PAD4), an enzyme critical for NETosis [14]. PAD4 hydrolyze peptidyl arginine into peptidyl citrulline on histones. Citrullination in histone proteins by PAD4 weakens the interaction of these proteins with negatively charged DNA and promotes heterochromatin decondensation and chromatin unfolding to form NETs [15].

A factor that is known to be critical for NETs formation and NETosis is the neutrophil elastase (NE) [4], that together with the release of neutrophil myeloperoxidase (MPO) from the granules translocate cleaves and digest histones to promote chromatin decondensation [5]. Taken together, MPO, NE and PAD4 are the major proteins involved in citrullination in histone and chromatin decondensation. These proteins are not only involved as mechanisms but also as markers of NETosis.

More than two decades ago, we have observed left ventricular hypertrophy (LV) and HF due to aortic stenosis is associated with three to four-fold increases in myocardial neutrophils as compared to control patients [6]. In animal models of HF, inflammatory infiltration of neutrophils was consistently found in the myocardial tissue suggesting that NETosis may promote the initiation and progression of HF [7].

The possible role of NETosis in myocarditis has recently emerged from studies in COVID-19 patients with “fulminant myocarditis” [8–10]. Although there are no direct data supporting the presence of NETosis in the heart, the presence of NETosis in lung and coronary artery autopsies suggest that COVID-19 activates NETosis, which lead to excessive tissue inflammation and thrombotic microangiopathy [11–14].

From this background, the present study aimed to investigate the occurrence of NETosis in the failing human heart due to cardiomyopathies (CMP) unrelated to COVID-19 infection. We have instigated and quantified the established markers of NETosis at the protein and mRNA levels and have proven for the first time that NETosis is an important component of chronic inflammation in patients with HF.

## Material and Methods

### Patients

Human LV samples were collected from the explanted hearts of patients undergoing orthotopic heart transplantation because of end-stage heart disease. All patients were severely symptomatic (NYHA grade II-IV) with poor LV systolic function. Patients either had normal coronary arteries with previously proven myocarditis and subsequently developed chronic HF with low ejection fraction (infCM, n = 7) or without histological evidence of myocardial inflammation (idiopathic dilated cardiomyopathy, DCM, n = 7), or had severe coronary artery disease (ischemic cardiomyopathy, ICM, n = 7) with a history of previous myocardial infarction (7 of 7). From the latter patients, only the regions distant from myocardial infarctions (remote regions) were investigated. Clinical data are summarized in Table. LV myocardium from 4 donor hearts that for technical reasons were not used for transplantation and intraoperative myocardial biopsies from one patient with atrial septal defect but with preserved LV function served as control tissues. All studies complied with the Declaration of Helsinki and were approved by the ethical committee of the Landesärztekammer Hessen, Frankfurt am Main (project numbers FF 8/2011 and FF 56/2012) and all patients gave written informed consent.

**Table 1.** Baseline characteristics of patients.

|  | Control | DCM | ICM | infCMP |
| --- | --- | --- | --- | --- |
| Number | 5 | 7 | 7 | 7 |
| Age | 51.4 ± 5.6 | 49.3 ± 6.7 | 55.2 ± 3.5 | 46.4 ± 4.49 |
| Sex<br>(male/female) | 4/1 | 6/1 | 6/1 | 4/2 |
| NYHA class, n | - | IV (7/) | IV (77) | IV (7/7) |
| EF (%) | >60% | 15 ± 2.1 | 21 ± 2.2 | 22 ± 4.1 |
| Hypertension | - | 1/7 | 4/7 | 2/7 |
| Diabetes | - | 2/7 | 4/7 | 3/7 |
| Chronic<br>pulmonary<br>disease | - | 2/7 | 2/7 | 3/7 |
| ACE inhibitors<br>(n) | 1 | 3 | 3 | 3 |
| Beta-blockers (n) | 1 | 2 | 2 | 3 |
| Ca2+ blockers<br>(n) | 1 | 2 | 2 | 3 |
| Digitalis (n) | - | 5 | 5 | 4 |
| Diuretics (n) | 1 | 6 | 6 | 5 |

### Tissue sampling

Myocardial tissues were taken postoperatively from the LV and washed with a cold Krebs–Henseleit solution augmented with 0.1% adenosine and 0.5% albumin. Tissue samples either were immediately frozen for immunohistochemistry and Western blot and stored at -80°C.

### Immunohistochemistry

The tissue samples were mounted in Tissue-TeK® O.C.T.TM (Sakura) and 5 µm thick cryosections were prepared using a Leica CM3050S cryotome. To maximally preserve NET structures, cryosections were mounted on negatively charged glass slides and fixed in 1% paraformaldehyde with 20% sucrose by -20^0^C as previously described [15]. Frozen sections were further fixed for 10 min with 4% paraformaldehyde at +4^0^C. After washing in phosphate buffered saline (PBS) sections were incubated with 1% bovine serum albumin for 30 minutes to block non-specific binding sites. and then incubated with the primary antibodies (Table S1) and were detected with anti-mouse or anti-rabbit IgG-conjugated with Cy3 or Cy2 (Biotrend). The nuclei were stained with 1 μg/ml 4′,6-diamidino-2-phenylindole (DAPI, Molecular Probes). F-actin was fluorescently stained using FITC-conjugated (Sigma) or Alexa633-conjugated phalloidin (Molecular Probes). Negative controls were obtained by omitting the primary antibody, in an otherwise similar protocol. Sections were embedded in Mowiol and coverslipped.

### Confocal microscopy

Tissue sections were examined by laser scanning confocal microscopy (Leica TCS SP2 and Leica SP5). Series of confocal optical sections were taken using a Leica Planapo x40/1.00 or x63/1.32 objective lens. Each recorded image was taken using four channel scanning and consisted of 1024 x 1024 pixels. To improve image quality and to obtain a high signal to noise ratio, each image from the series was signal-averaged. After data acquisition, the images were further processed for restoration, quantification and three-dimensional reconstruction using an Imaris multichannel image processing software (Bitplane, Zürich, Switzerland).

### Quantitative immunofluorescent microscopy

Immunofluorescence measurements were done using a x40 Planapo objective (Leica) and a Leica (Leitz DMRB) fluorescent microscope equipped with a Leica DC380 digital camera. Cryosections from at least two different tissue blocks in each case were used. For quantitative analysis all sections were immunolabeled simultaneously using identical dilutions of primary and secondary antibodies and other reagents. Immunofluorescent images were obtained under identical parameters of imaging, zoom, pinholes, objectives, and fluorescence power. Sections exposed to PBS instead of primary antibodies served as negative controls. The image acquisition settings were standardized for all groups to ensure that the image collected demonstrated a full range of fluorescence intensity from 0 to 255 pixel intensity level and were kept constant during all measurements. Quantification of the studied proteins was performed blindly, having on the screen only one channel showing F-actin labelling. For each patient at least 10 random fields of vision were analyzed using image analysis software (Leica) and Image J program as described. Arbitrary units were calculated per unit surface area (AU/100 μm2 or AU/1 mm^2^).

### Western blot analysis

Tissue samples were processed for Western blot analysis as previously described [35, 36]. In brief, frozen tissues were homogenized in RIPA buffer (containing 20 mmol/L Tris–HCl at pH 7.4, 100 mmol/l NaCl, 5 mmol/L thylene-diamine tetra-acetic acid, 1% Triton X-100, 10% glycerol, 0.1% sodium dodecylsulfate, 1% deoxycholate, 50 mmol/L NaF, 10 mmol/L Na4P2O7, 1 mmol/l Na3VO4, 1 mmol/L phenylmethylsulfonylfluoride and mammalian protease inhibitor cocktail (Sigma) at pH 7.4 and centrifuged at 2000 ×g at 4 °C for 10 min. Cellular and LV myocardial extracts were loaded onto 12% polyacrylamide gel and separated under the reducing conditions. Proteins were electro transferred onto nitrocellulose membrane (Invitrogen) and blocked with 5% non-fat dry milk in Tris-buffered saline Tween-20 (TBST) at 4 °C. After washing with TBST, proteins were exposed overnight at 4°C to primary antibodies (Table S1) diluted in TBS with 5% powdered milk. Bound antibodies were detected by peroxidase-conjugated anti-mouse IgG horseradish peroxidase-conjugated and SuperSignal WestFemto (Pierce) detection system and exposed to X-ray film. Quantification of immunoblots was done by scanning on a STORM 860 (Amersham, Pharmacia Biotech) using ImageQuant software as described [36]. The immunoblotting values for the investigated proteins were normalized per beta-actin (Sigma). The control values were set at 100% (see Figure 7, as an example).

### Quantitative Real Time PCR

RNA was isolated from 30 mg of ventricular tissue with a RNeasy mini tissue kit (Qiagen). cDNA libraries were synthetized on the total RNA using High Capacity cDNA reverse transcriptase kit (Applied Biosystems). The list of PCR primer sets is listed in Table S2. For quantitative PCRs, the cDNA pool was diluted 1 in 10 with miliQ water and used as a template. To assess expression of gene levels, the quantitative PCR was performed with 25-35 cycles using a standard PCR thermocycler (BioRad laboratories). GAPDH was served as a reference gene. Both transcripts were detected using on the 1% agarose gel developed at 5V/cm2.

### Statistical analysis

All statistical analyses were performed using SPSS 25.0 program for Mac. The results are given as median with 25th/50th/75th quartile (interquartile range, IQR) for continuous variables. For multiple comparisons, we used ANOVA followed by an analysis with the Bonferroni t-test. Associations between variables were assessed using Spearman’s rank correlation. A P value of <0.05 was deemed statistically significant.

## 3. Results

### The number of neutrophils and macrophages is drastically increased in HF

We used immunolabeling to identify the myocardial neutrophils. Figure 1 show typical confocal images of neutrophils in control patients and in patients with HF. It is worth mentioning that in 3 of 7 patients with inflCMP and in 1 of 7 ICM patients foci of neutrophil accumulations have been detected (Figure 2A and 2B). Figure 2B shows the quantification of the number of neutrophils per myocardial surface and shows a 3 to 4-fold increase in the number of neutrophils in CMP hearts compared to control hearts. It was found that 1 mm^2^ of the myocardial surface comprised an average of (2.2-4.4) neutrophils in the control group, 14.2 (12.9-17.1) in DCM, 17.4 (14.5-19.9) in ICM and 19.3 (17.8-22.6) in infCMP. It should be emphasized that the number of neutrophils in the latter group differed significantly from the number of neutrophils in the ICM (p<0.01) and DCM (p<0.01) groups. Similar results were obtained using an anti-CD66b antibody (Figure S1A and S1B).

**Figure 1.**
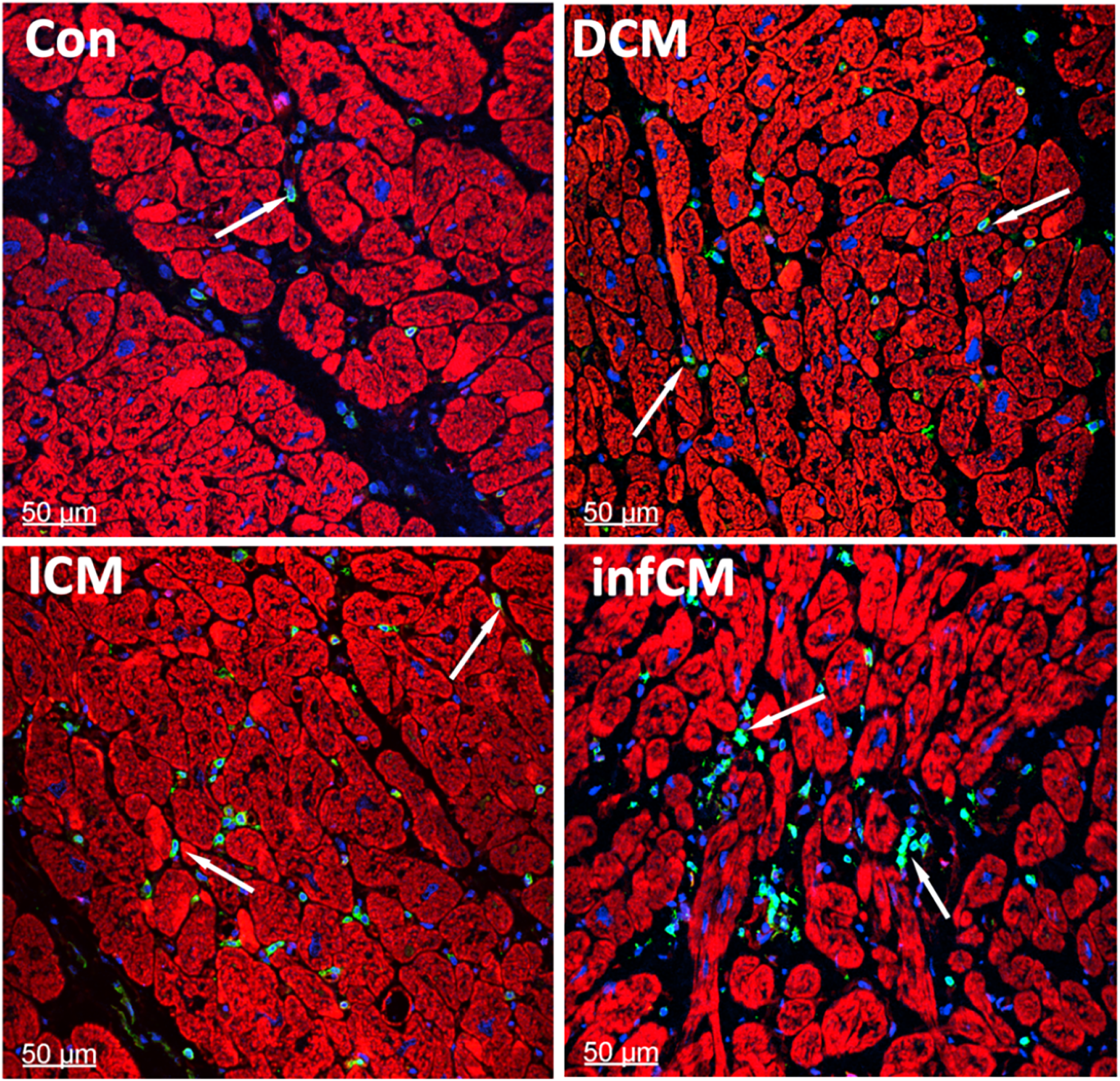
Neutrophils are increased in HF patients. **A**, Representative confocal micrographs of CD45-positive cell (green), arrows) in control (A), DCM (B), ICM (C) and inflCMP (D). Notice a significant increase in CD45-positive cells in CMP patients. Nuclei are shown in blue after staining with DAPI and F-actin is shown in red after labelling with Alexa633-phalloidin.

**Figure 2.**
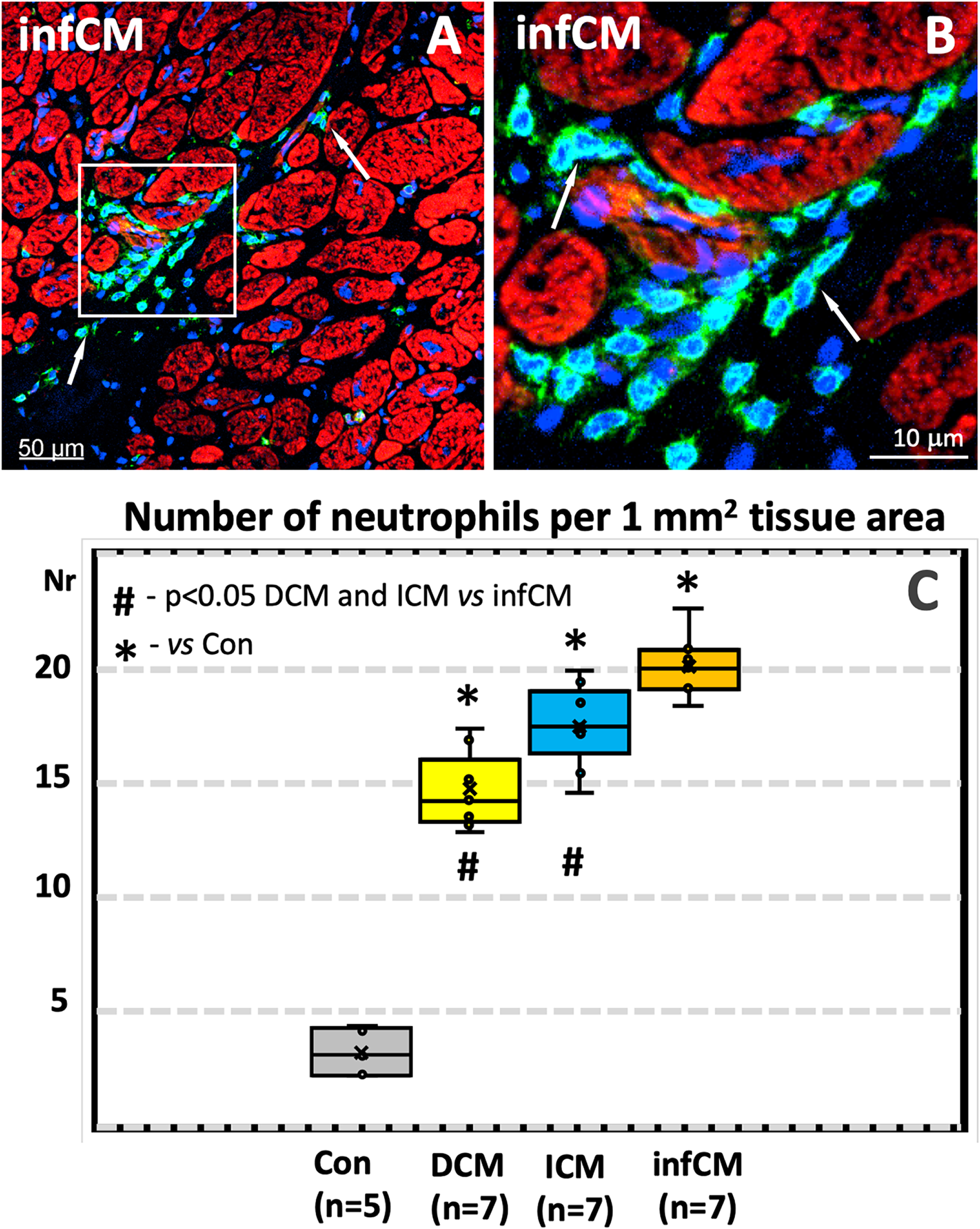
Focal accumulations of neutrophils and increased number of neutrophils in HF due to CMP. **A,** A typical example of neutrophil accumulations in a patient with inflCMP, which are shown at higher magnifications (**B**). **C**, Quantitative analysis of CD-45-positive cells in the studied groups. Values are presented as median with IQR.

We used a CD68 antibody to quantify the number of macrophages and the results are presented in Figure S2A. It was found that 1 mm^2^ of the myocardial surface in the control group comprised an average of 13.3 (7.3-18.5) macrophages. The same ventricular myocardial surface comprised 49.4 (42.9-61.5) macrophages in DCM, 57.7 (48.5-65.9) in ICM and 58.5 (56.4-66.9) in infCMP. The difference between all HF groups and control was statistically different (p<0.01) The difference between DCM and infCMP was statistically different (p<0.01). The increase in the number of macrophages indicates the presence of chronic myocardial inflammation in HF patients. In addition, the increase in the number of macrophages correlated strongly (r = 0.918, p<0.01) with the number of neutrophils (Figure S2B).

### The majority of neutrophils undergoes NETosis in patients with HF

We used confocal microscopy and immunohistochemistry with antibodies against neutrophils and NETosis to study the proportion of neutrophils undergoing NETosis. Figure 3 shows that MPO- and citH3-positive cells are obviously increased in HF patients as compared to control patients. Figure S3A and S3B show that PAD4 is clearly confined to the neutrophil nuclei. Figure 4A and 4B show typical MPO- and NE-positive NETosis cells. The quantification of the proportion of neutrophils undergoing NETosis is shown in Figure 4C. In patients with inflCMP, the median percentage of neutrophils undergoing NETosis was 49.4% (44.9-52.1). In patients with ICM and DCM, the median percentage of neutrophils that underwent NETosis was 45.3% (37.9.9-46.1) and 41.5% (37.2-47.4), respectively. The difference in the number of neutrophils with NETosis in patients with inflCMP was statistically significant compared to ICM (p<0.05) and DCM (p<0.01). In control patients, less than 4 % neutrophils with positive NETosis markers were found. Similar results were obtained using citH3 and NE as markers of NETosis (Figure S4).

**Figure 3.**
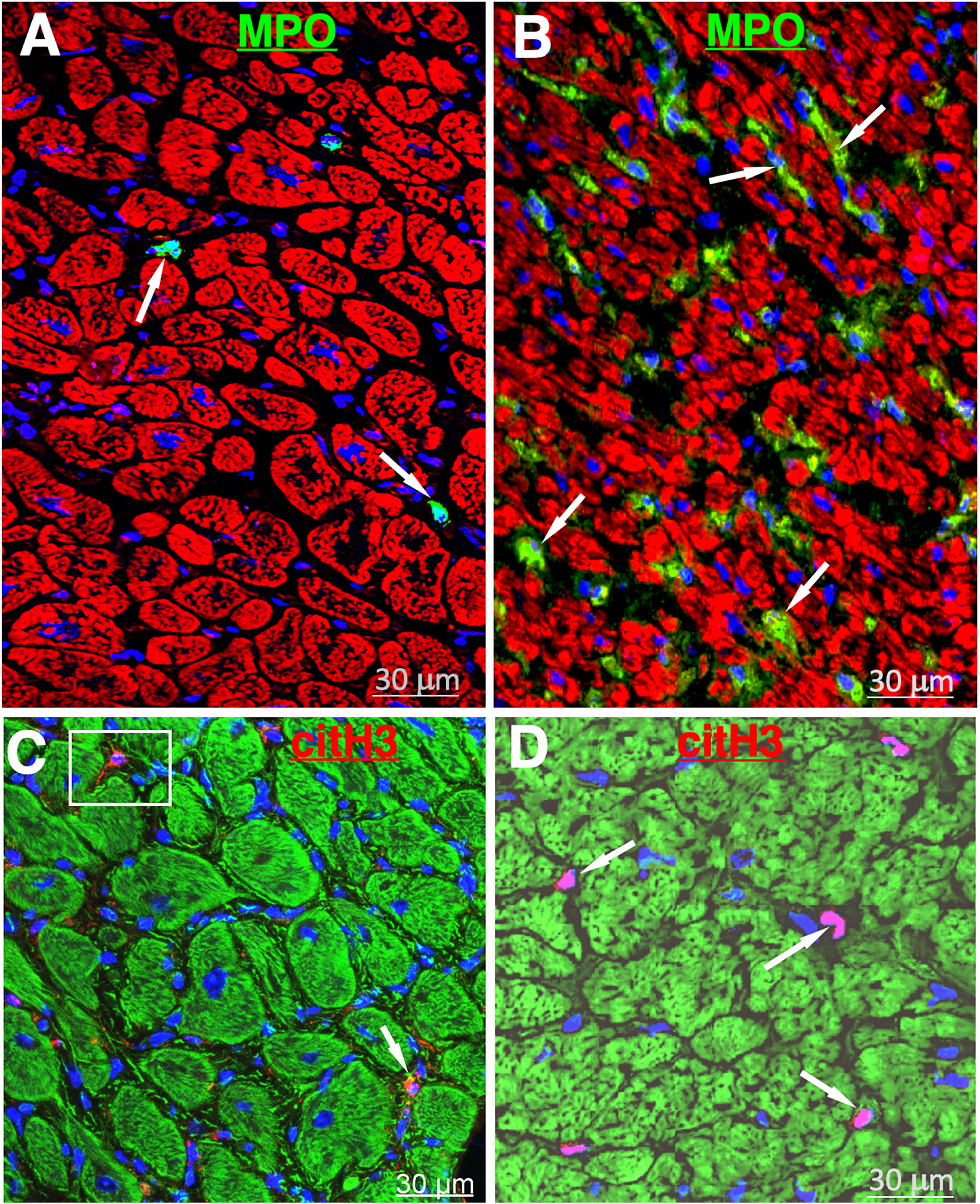
MPO and citH3 are increased in HF patients. **A**, MPO expression (green, arrows) in a control (**A**) and a patient with DCM (**B**). CitH3 expression (red), arrows) in a control (**C**) and a patient with DCM (**D**). Notice a significant increase in both proteins in HF patients. Nuclei are shown in blue and F-actin is shown in red (A) after labelling with Alexa633-phalloidin or green labelling with Cy2-phalloidin.

**Figure 4.**
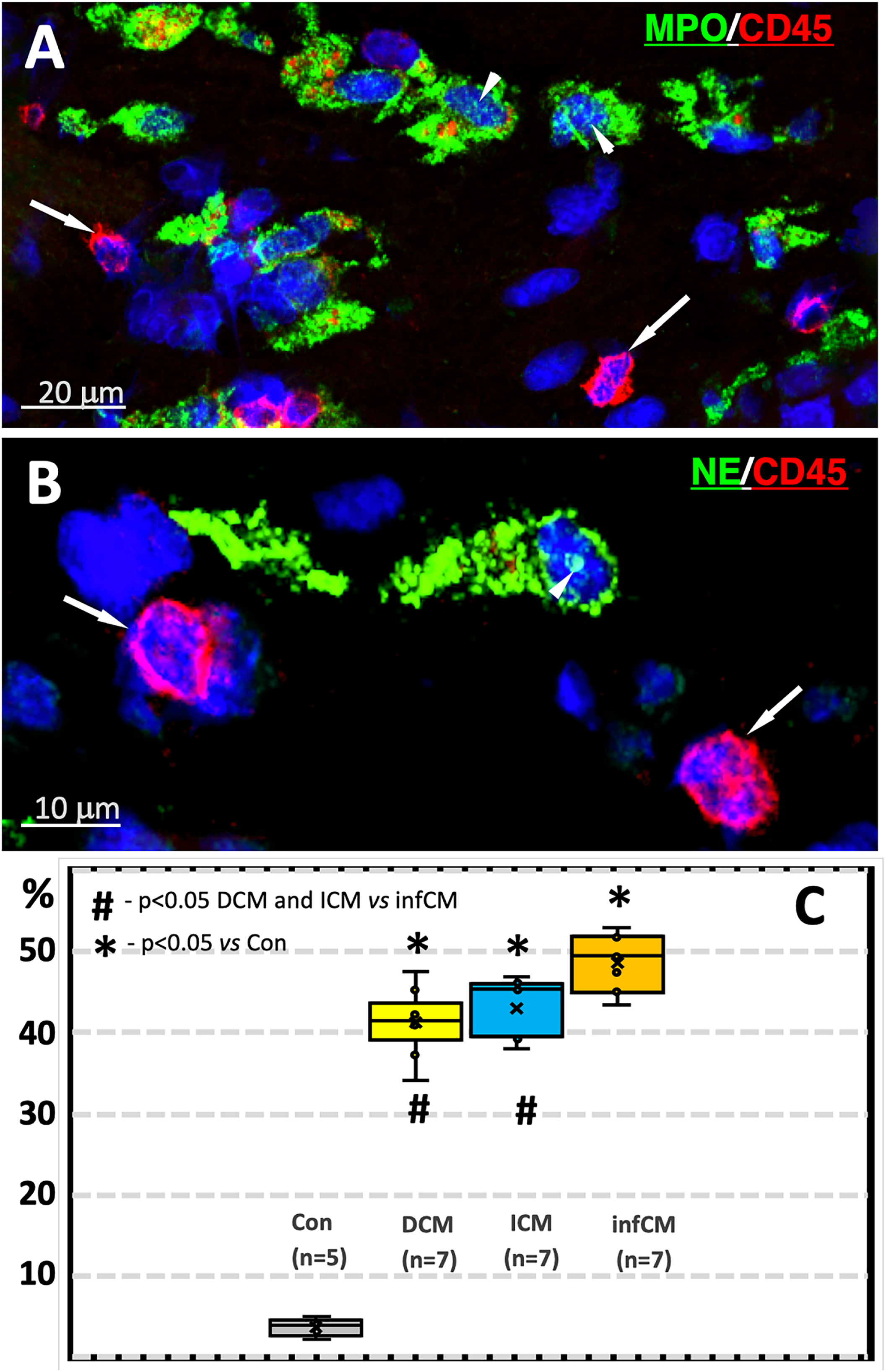
Neutrophils undergoing NETosis are increased in HF patients. Representative confocal images of double labeling of MPO (green) with CD45 (red) and NE (green) wtih CD45 (red) in a patient with inflCMP (**A**) and in a patient with ICM (**B**). Arrows denote intact, naïve neutrophils and arrowheads indicate the presence of both proteins in the nucleus of neutrophils. Nuclei are shown in blue after staining with DAPI. **C**, Quantification of the percent of neutrophils undergoing NETosis using MPO immunolabeling. Values are presented as median with IQR.

Next, we used MPO/citH3 and NE/citH3 dual labeling protocols. Confocal photomicrographs in Figure 5A and 5B show that these two markers are colocalized in most NETosis cells. However, there are cells in which these two markers are not colocalized. The quantification of NETosis using citH3/NE immunolabeling is shown in Figure 5C. In patients with inflCMP, the median percentage of neutrophils positive for both, citH3 and NE was 79.4% (71.1-84.9). In patients with ICM and DCM, the mean percentage of citH3 and NE positive neutrophils was 57.9% (52.2-64.8) and 55.6% (47.2-60.9), respectively. The difference between inflCMP was statistically significant compared to ICM (p<0.01) and DCM (p<0.01). In control patients, less than 4% (2.9-4.9) neutrophils were found to be positive for citH3/NE markers.

**Figure 5.**
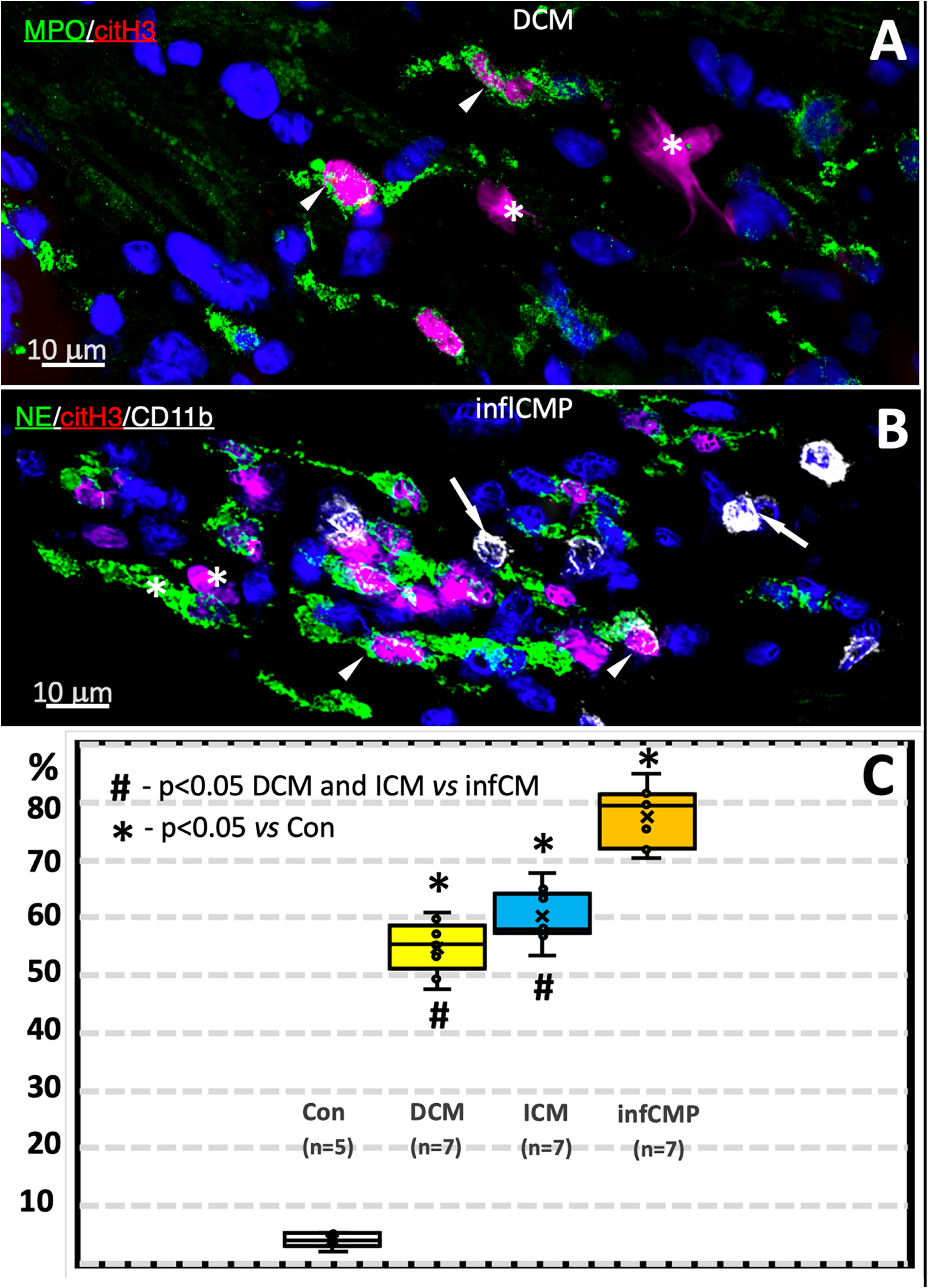
The use of dual markers increases the detection rate of NETosis in HF patients. **A**, Representative confocal image of double labeling of MPO (green) with citH3 (red) in patients with DCM (**A**) or NE (green) with citH3 (red) in a patient with inflCMP (**B**). In both panels, arrowheads denote co-localization of these NETosis markers, while asterisk indicate the absence of their co-localization. Arrows in (**B**) indicate intact CD11b cells. **C**, Quantitification of Netosis using double labeling for citH3 and NE. Values are presented as median with IQR. Notice that compared with double labeling with 2 NETsis markers (this Figure) considerably increases the percent of NETosis detection as compared with single labeling (Figure 4).

### NETs structures are markedly increased in HF patients

An important function of neutrophil is the formation of extracellular NETs. We have studied NETs in ventricular tissue *in situ* using confocal microscopy and immunohistochemistry with antibodies against NETosis. Figure 6A, 6B and 6C demonstrate typical NETs as long (range 5 μm – 50 μm), very thin structures in a granular or fibrillar pattern. Quantification of NETs using double labelings is presented in Figure 6D and 6E. In patients with inflCMP, the median percentage of neutrophils that developed citH3/NE-positive NET structures relative to the total number of neutrophils was 11.6 (8.6-18.9). In patients with DCM and ICM, the median number of neutrophils that developed NETs was 10.3 (8.6-17.4) and 11.7 (8.78-16.4), respectively. The difference between the HF groups was not statistically significant. Similar results were observed when citH3/MPO-positive NETS structures were quantified (Figure 6E). It is worth noting that our results showed that the number of neutrophils undergoing NETosis was almost 5-fold higher than that of neutrophils in NETosis developing NETs structures (compare Figure 5C with Figure 6D and 6E).

**Figure 6.**
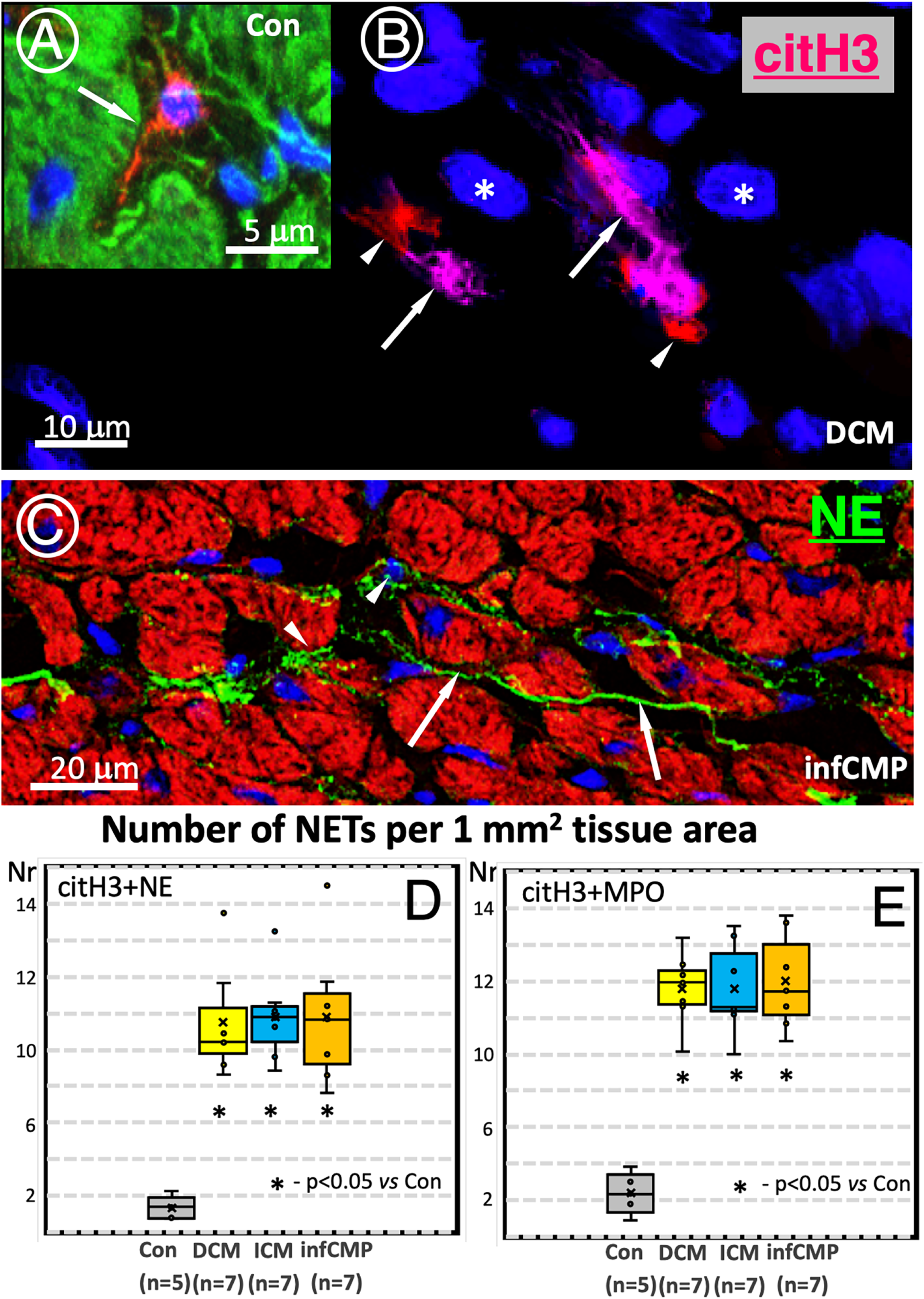
Examples of NETs and increased NETs number in HF patients. Typical confocal images of citH3 (red) forming long interstitial structures (arrows) in a control (**A**) and in a patient with DCM (**B**). Asterisks indicate citH3 nuclei and arrowheads denote citH3 that is not co-localized with DNA-DAPI (**B**). **C**, NE-positive extracellular NET structures in a long granular or fibrillar patterns (arrows) in a patient with inflCMP. Arrowheads denote perinuclear NE-positive granules in probably intact neutrophils. Nuclei are shown in blue and F-actin is shown in green (**A**) after labelling with Cy2-phalloidin or red (**C**) after labeling with Alexa633-phalloidin. Quantification of NETs in the studied groups using double labeling for citH3 and NE (**D**) or citH3 and MPO (**E**).

### The proteins and genes involved in NETosis are significantly increased in HF

We performed Western blot and quantitative analysis of the fluorescence intensity of citH3, MPO, NE and PAD4. Figure 7A show a representative Western blot gel of citH3 and its quantification is presented in Figure 7B. The results showed that the citH3 protein level in patients with inflCMP was 4.8-fold higher compared to control (p<0.01). In patients with ICM and DCM, citH3 was 4.4-fold (p<0.01) and 2.7-fold (p<0.01), respectively higher than in control. The difference in citH3 expression in DCM patients was statistically different from patients with ICM (p<0.01) and inflCMP (p<0.01). Measurements of the PAD immunofluorescence intensity showed that all HF groups differed significantly from the controls (p<0.01). The difference between HF groups was not statistically significant (Figure 7C). Similarly, measurements of the MPO (Figure 7D) and NE (Figure 7E) immunofluorescence intensity showed that all HF groups differed significantly from the controls (p<0.01). However, the MPO fluorescence intensity values in patients with infCMP was statistically significant as compared with ICM (p<0.01) and DCM (p<0.01 (Figure 7D). In contrast, the NE fluorescence intensity values in patients with infCMP differed statistically only in comparison to ICM patients (p < 0.01).

**Figure 7.**
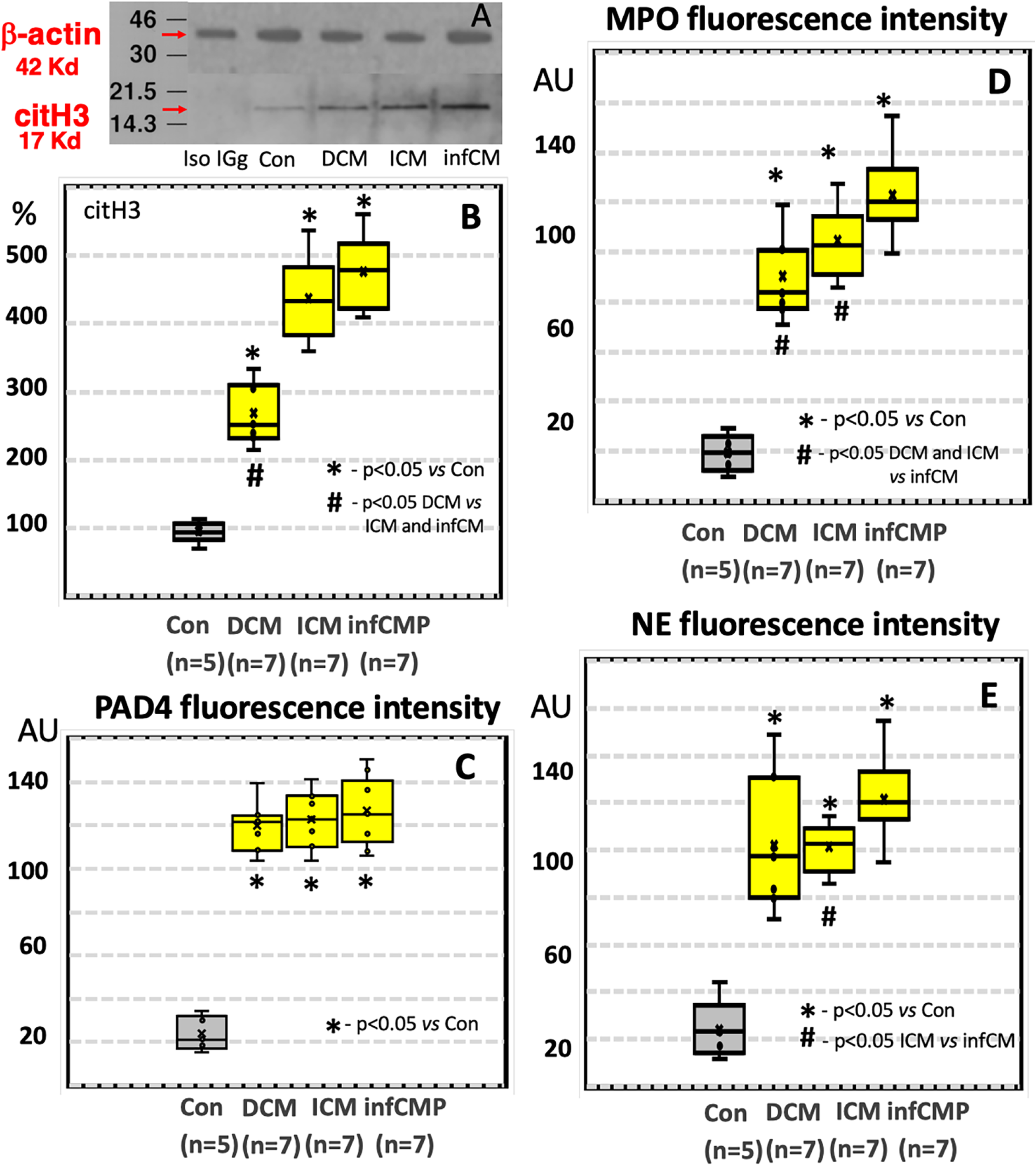
CitH3, PAD4, MPO and NE proteins are increased in HF patients. Representative Western blot of citH3 (**A**) and its densitometric quantification (**B**). Quantification of the fluorescence intensity of PAD4 per 100 μm^2^ nuclear area (**C**) and MPO (**D**) or NE (**E**) per 1 mm^2^ myocardial area. Values are presented as median with IQR.

Next, we performed quantitative PCR analysis of the genes involved in NETosis. We found that the mRNA levels of MPO (Figure S5A), NE (Figure S5B) and PAD4 (Figure S6) were 3- to 4-fold higher in the HF groups compared with the control group (p<0.01).

## Discussion

In this study, we show for the first time an increased number of neutrophils and the presence of NETosis in the ventricular myocardium in patients with HF due to CMP, regardless of its etiology. This is important to understand that NETosis contributes to the pathophysiology of HF.Importantly, using two markers for NETose increases their detection by 40% (compare Figure 4C with Figure 5C).

It is noteworthy that NET structures are very fragile structures and that it is difficult to recognize them *in situ* compared to NETs structures in neutrophils *in vitro*. For this reason, we found that the number of neutrophils with NET structures was several times lower than the number of neutrophils that underwent NETosis. Nevertheless, the number of NETs was three to four times higher in HF patients compared to the control group.

The clinical significance of our findings lies in the fact that increased levels of NETosis may increase tissue damage and further sustain chronic myocardial inflammation in HF and hence may be a potential target for future therapeutic interventions [16–18].

At present, there are different approaches to treat NETosis. One of these approaches is to prevent excessive neutrophil activation, neutrophil degranulation and inhibit the components involved in the NETosis program (PAD4, MPO and NE). The present study demonstrated significant increases in HF of all these components.

According to recent genetic and pharmacological studies, MPO, NE and PAD4 inhibitors are effective in adverse post-myocardial infarction remodeling, DCM, decompensated ventricular hypertrophy and age-related myocardial fibrosis whose deterioration lead to HF [19–23].

Another approach is based on the destruction of NETosis or the attenuation of its harmful effects on the heart. The most potent effect of the latter processes is exerted by recombinant human DNase (dornase-α), also known as pulmozyme, which was developed to degrade the large amount of free extracellular DNA fragments and promote clearance of NET formation. It is commonly used in patients with cystic fibrosis. The improvement in lung function and low risk of side effects indicate a significant net benefit of dornase-α [24]. Therefore, it is tempting to speculate the positive effects of the dornase-α also in patients with HF.

Experiments using neutrophils *in vitro* have shown that NETosis can be prevented by microtubule inhibitor colchicine [25]. A recent experimental study using a high salt diet-induced HF in rats showed that colchicine attenuates cardiac inflammation and fibrosis [26]. In addition, clinical trials testing efficacy of colchicine against diverse cardiovascular diseases including HF showed promising beneficial effects [27]. It should, however, be mentioned that the effects of colchicine in patients with chronic HF is limited to a single randomized controlled trial showing that colchicine although effective in reducing inflammation biomarker levels, did not affect NYHA class or the likelihood of death or hospital stay for HF [28]. In addition, the *in vitro* and *in vivo* experiments have shown that simvastatin and metformin can inhibit the formation of NETs and NETosis [29, 30]. Several recent clinical studies have shown that statins and metformin, owing to their anti-inflammatory effects, may effectively improve HF [31, 32]. For example, rosuvastatin, significantly decreases the plasma MPO level and the number of circulating neutrophils in patients with chronic HF [33]. Similarly, metformin also decreased plasma MPO levels and prevents myocardial neutrophil infiltration and activation in rats with isoproterenol-induced HF [34].

Taken together, these experimental, clinical studies and our observations indicate that inhibiting NETosis might be beneficial for improving HF.

### Study limitations

The findings of the present study should be interpreted in light of certain limitations. The major limitation is the small number of the investigated patients resulting in the high interindividual variability of the parameters studied. Nonetheless, our study demonstrating dramatic increases in neutrophils undergoing NETosis and the major proteins and genes involved in NETosis justify our conclusions.

## Conclusions

This is the first study to show the occurrence of NETosis in human hearts in situ and demonstrate that NETosis is an important component of the low-grade myocardial inflammation in HF.

## Data Availability

The data underlying this article will be shared on reasonable request to the corresponding author

## Acknowledgments

The authors thank Brigitte Matzke, Beate Grohmann and Dr. Viktoria Polyakova for excellent technical assistance. This work was supported by the Stiftung William G. Kerckhoff-Herz-und Rheumazentrum Bad Nauheim (in support of M.R.).

## Nonstandard Abbreviations and Acronyms

CMP: cardiomyopathy
citH3: citrullinated histone H3
DCM: dilated cardiomyopathy
HF: heart failure
ICM: ischemic cardiomyopathy
inflCMP: inflammatory cardiomyopathy
IQR: interquartile range
MPO: myeloperoxidase
NE: neutrophil elastase
NET: neutrophil extracellular trap
NETosis: NET-dependent death of neutrophils
PAD4: peptidyl arginine deiminase

qPCR

